# Between-centre differences for COVID-19 ICU mortality from early data in England

**DOI:** 10.1101/2020.04.19.20070722

**Authors:** Zhaozhi Qian, Ahmed M. Alaa, Mihaela van der Schaar, Ari Ercole

## Abstract

The high numbers of COVID-19 patients developing severe respiratory failure has placed exceptional demands on ICU capacity around the world. Understanding the determinants of ICU mortality is important for surge planning and shared decision making. We used early data from the COVID-19 Hospitalisation in England Surveillance System (from the start of data collection 8^th^ February -22^nd^ May 2020) to look for factors associated with ICU outcome in the hope that information from such timely analysis may be actionable before the outbreak peak. Immunosuppressive disease, chronic cardiorespiratory/renal disease and age were key determinants of ICU mortality in a proportional hazards mixed effects model. However variation in site-stratified random effects were comparable in magnitude suggesting substantial between-centre variability in mortality. Notwithstanding possible ascertainment and lead-time effects, these early results motivate comparative effectiveness research to understand the origin of such differences and optimise surge ICU provision.

## Introduction

Since the first cases in November 2019, the spread of SARS-CoV-2 infections has placed unprecedented strain on healthcare. The intensive care unit (ICU) is of particular concern as large numbers of patients with severe respiratory complications mean that in some areas, ICUs have been completely overwhelmed.

Understanding the determinants of ICU outcome is crucial both for surge planning and shared decision making. Whilst a number of risk scores have been published [1] they do not specifically look at this population. Furthermore, ICU availability, admission policy and structure varies across Europe [2] as do demographics and government policy. Thus, it is likely that ICU outcomes could also vary significantly by region motivating an individualised modelling approach. UK mortality has been particularly high and we sought to urgently identify predictors of mortality in patients admitted to the ICU with COVID-19 [3].

## Data and analysis

We obtained de-identified COVID-19 Hospitalisation in England Surveillance System (CHESS) data from Public Health England (PHE) for the period from 8^th^ February (data collection start) to 22^nd^ May 2020 (5,062 cases ICU cases– 1,547 deaths, 1,618 discharges from 94 NHS trusts across England). Mean APACHE-II score for each site from ICU national audit data for COVID-19 patients over a similar period was used to attempt to correct for case-severity/as a proxy for admission policy since a variety of presentation severity indicators may also drive outcome [4].

We used a Cox proportional hazards mixed-effects model for mortality, with NHS trust as the random effect. The estimated coefficients associated with each predictor are shown in Figure 1. The proportional hazards assumption was not violated (*p* = 0.061).

**Figure 1:**
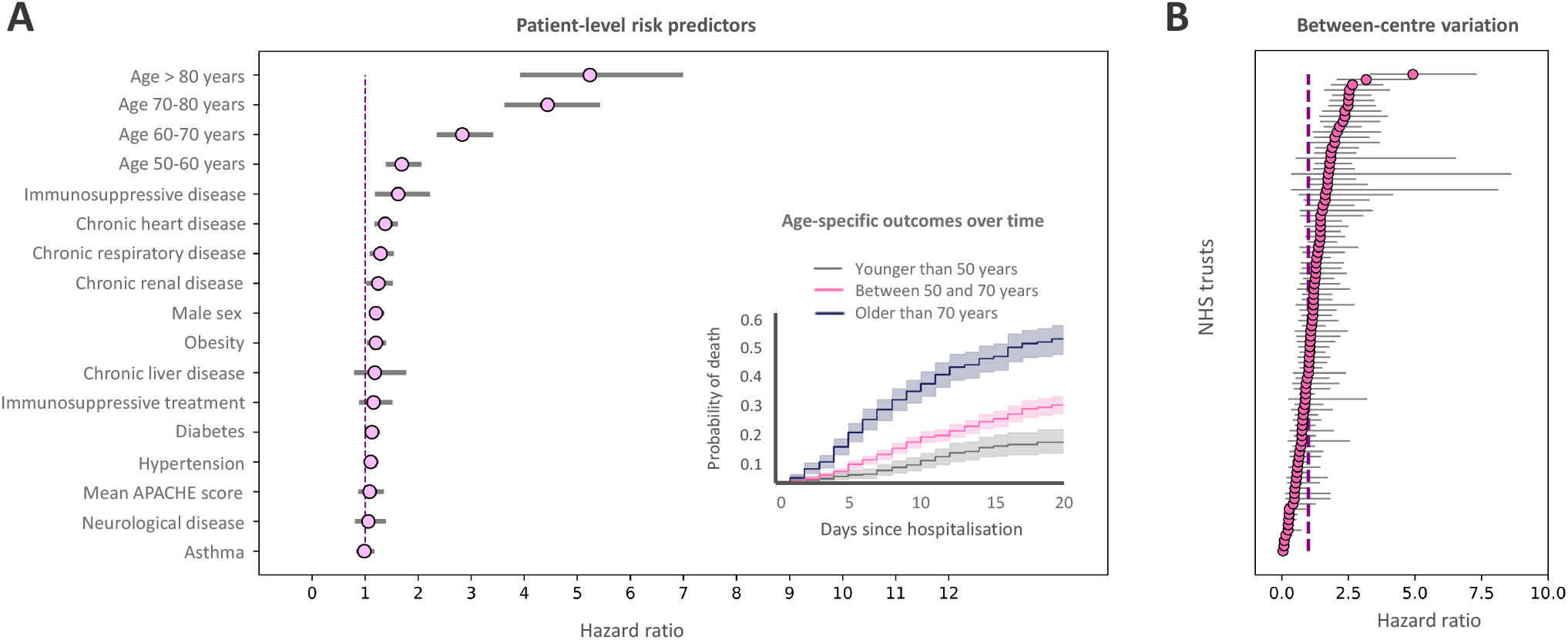
***Panel (A)*** *Figure 1: Panel (A) Fixed effects estimates for CHESS predictors and mean APACHE-II score for each site from national audit data. The strongest patient-factor predictors are older age, immunosuppressive disease and chronic heart / renal disease. The inset shows mortality probability over time for young, intermediate and older age groups*. ***Panel (B)*** *random effects showing between centres variation showing a hazard ratio variation between sites from 0 to over +4 which is comparable in magnitude to the strongest CHESS predictor (older age) showing that between-centres variation is an appreciable determinant of outcome. 95% CIs shown*.

## Discussion and conclusions

Immunosuppression, chronic heart/renal disease and age were key predictors of mortality. In comparison with these fixed effects, the magnitude of the between-centre variation (hazard ratio between 0 to over +4) is comparable to the strongest fixed effects predictor. The cause of such between-centre variation is unclear and may have a variety of residual case-mix or structural explanations. In particular, ICU demand varies both regionally and locally and we may hypothesize that high levels of strain or constraints on surge capacity could be actionable determinants, although we do not have data to examine this. Such considerations are important to understand as they may influence optimal configuration or transfer considerations locally.

Analysis limitations include possible incomplete ascertainment (particularly before approximately 15^th^ March), potential lead time bias from earlier deaths in the elderly group (although a sensitivity analysis excluding patients with < 7 days follow-up yielded qualitatively similar results-Supplement) and an inability to track patient transfers. Nevertheless, the magnitude of the random effects is striking. This motivates urgent comparative effectiveness research to characterise between-centre differences to inform surge best-practice in both in England and elsewhere.

## Data Availability

Source data controlled by Public Health England. Under the terms of our data sharing agreement, the authors are not able to re-share the source data but will entertain re-quests for scientific collaboration.

## Acknowledgements

We would like to acknowledge PHE for providing us access to the CHESS dataset and the intensive care national audit and research centre for providing aggregate APACHE-II data, and also thank Anees Pari and Geraldine Linehan for their support.

## Availability of data and materials statement

Source data controlled by Public Health England. Under the terms of our data sharing agreement, the authors are not able to re-share the source data but will entertain requests for scientific collaboration.

## Competing interests

None to declare.

## Funding

Not externally funded.

## Authors’ contributions

AE and MvdS conceived of and supervised the study. ZQ and AMA coded the statistical models. All authors contributed to the manuscript.

## Notes

### Competing Interest Statement

The authors have declared no competing interest.

